# Vaccination status and re-infection among COVID patients admitted in COVID High Dependency Unit (HDU) of a tertiary hospital in Eastern Nepal: A cross-sectional study

**DOI:** 10.1101/2023.08.30.23294839

**Authors:** Sujan Kafle, Varsha Chhetri, Joshan Lal Bajracharya, Lenish Pokharel, Suyash Dawadi, Ujwal Basnet, Shreya Dhungana, Durga Neupane

## Abstract

**Introduction:** The relationship between COVID-19 vaccination and Coronavirus disease severity and outcomes remain topics of significant interest. This cross-sectional study done among COVID-19 patients admitted to the High-dependency unit of a tertiary hospital in Eastern Nepal aimed to assess the association between vaccination status, prior infection, and disease outcomes, and the modification of these associations by the presence or absence of comorbidities.

**Methodology:** Demographic and clinical data were collected from 102 COVID-19 patients admitted to the High-Dependency Unit of Mechi Zonal Hospital, including information on vaccination status, comorbidities, disease severity, and outcomes. Statistical analysis, including chi-square tests and Fisher’s exact tests, was performed to examine the associations.

**Results:** Among the study participants, 49% had received at least one dose of COVID-19 vaccine. Vaccinated individuals had a significantly lower rate of severe disease compared to non-vaccinated individuals (χ²=10.05, p=0.002). Recovery and mortality rates did not differ significantly between the two groups (χ²=1.008, p=0.315). However, when stratified by comorbidities, vaccinated individuals with comorbidities had higher recovery rates compared to non-vaccinated individuals (85.29% vaccinated vs. 25.00% non-vaccinated, Fisher’s exact test p=0.024). Vaccinated individuals, both with and without comorbidities, had lower rates of severe disease compared to non-vaccinated individuals. However, the association was found to be significant only in individuals with comorbidities (12.50% vaccinated without comorbidities vs. 47.92% non-vaccinated, p=0.017; 23.53% vaccinated with comorbidities vs. 75.00% non-vaccinated, p=0.065).

**Conclusion:** Our findings suggest that COVID-19 vaccination is associated with a reduced risk of severe disease among individuals with or without comorbidities and decreased risk of mortality among those with comorbidities. However, larger studies are needed to validate these findings and further explore the impact of vaccination on disease outcomes. These findings support the ongoing efforts to promote COVID-19 vaccination as a crucial public health intervention.

## Introduction

The novel coronavirus disease, COVID-19, has imposed significant challenge for public health officials and healthcare providers. According to the World Health Organization, over 6.9 million deaths have been implicated to COVID-19.^1^ Many questions are yet to be explored and answered regarding the virus, SARS-CoV-2. Much is understood about the viral infection and the modes of transmission, but the timing, magnitude, and longevity of humoral immunity after natural infection and vaccination are still a subject of study. ^2–4^

A breakthrough infection is an infection with a virus, bacterium or other germ after you have been vaccinated.^5^ Vaccine breakthrough infections are expected. COVID-19 vaccines are effective at preventing most infections. However, like other vaccines, they are not 100% effective.^6^ Fully vaccinated people with a vaccine breakthrough infection are less likely to develop serious illness than those who are unvaccinated and get COVID-19.^6^

For epidemiologic purposes, severe Covid-19 in adults is defined as dyspnea, a respiratory rate of 30 or more breaths per minute, a blood oxygen saturation of 93% or less, a ratio of the partial pressure of arterial oxygen to the fraction of inspired oxygen (PaO2:FIO2) of less than 300 mm Hg, or infiltrates in more than 50% of the lung field.^7,8^

The recommended dosage of AstraZeneca (ChAdOx1-S [recombinant] vaccine) is two doses given intramuscularly (0.5ml each) with an interval of 8 to 12 weeks.^9,10^ WHO recommends two doses of Janssen Ad26.COV2.S COVID-19 vaccine. The second dose should be given 2-6 months after the first dose.^11^ The recommended dose of Sinopharm COVID-19 vaccine (Verocell) is 2 doses (0.5 mL each) at a recommended interval of 3 to 4 weeks.^12^

We conducted this study to try and attempt to answer few questions – first, among the hospitalized participants, whether there was a significant difference in disease severity and mortality among the vaccinated and non-vaccinated particpants; second, whether there was a significant difference between disease severity and mortality among reinfected individuals and new cases.

In summary, the aim of this study is to evaluate the effectiveness of vaccination in preventing severe disease and mortality in individuals with and without comorbidities. We aim to assess the impact of vaccination on the adverse outcomes, including severe disease and mortality by analyzing the vaccination status of participants who experienced such outcomes. We believe that the findings of this study will contribute to our understanding of the role of vaccination in mitigating the severity of illness and reducing mortality, particularly in individuals with underlying comorbidities. This knowledge can inform public health strategies and guide decision-making regarding vaccination recommendations for vulnerable populations.

## Material and Methods

The required information and data for this study were collected from various sources like PubMed, World Health Organization (WHO), Johns Hopkins Medicine, Center for Disease Control (CDC) and the New England Journal of Medicine (NEJM). The key terms used were COVID-19, coronavirus, SARS-CoV-2, breakthrough infection, vaccination, and severe COVID-19.

A descriptive cross-sectional study was carried out among the cases of COVID-19 admitted in the High Dependency Unit (HDU) of Mechi Zonal Hospital, Nepal for two months (January 26^th^ to March 26^th^, 2022). The phone number of the participants or their guardians were obtained from the registry of the HDU. The participants or their guardians (in case of the death of the participant) were contacted via telephone and the information sheet was read out to them in the Nepali language (which they preferred). The participants (or their guardians, in case of the participant’s death) who provided verbal consent for the study were enrolled in the study.

The ethical clearance was obtained from the Nepal Health Research Council on April 17^th^, 2022. The study was started on June 1^st^, 2022, and the data collection was completed on June 18^th^, 2022. We used a pre-tested questionnaire for conducting this study. With the help of the questionnaire, data were collected from the participants or their guardians. The information about the hospital stay of the participants was obtained from the medical records section of the hospital.

The data was collected, tabulated, and statistically analyzed using Statistical Package for Social Sciences version 25.

## Results

During the study period, there were a total of 120 cases of COVID-19 admitted in the High Dependency Unit of Mechi Zonal Hospital, Nepal. 18 participants or their guardians either declined to participate or couldn’t be contacted. So, we collected the data from 102 participants or their guardians for the study.

Among the study participants, 52% were male and 48% were female. The mean age of our study population was 48.34 ± 26.15 years with a minimum age of 11 days and a maximum age of 96 years. The age category with the highest representation was 18-59 years, comprising 43.14% of the participants (Figure 1).

**Figure 1:**
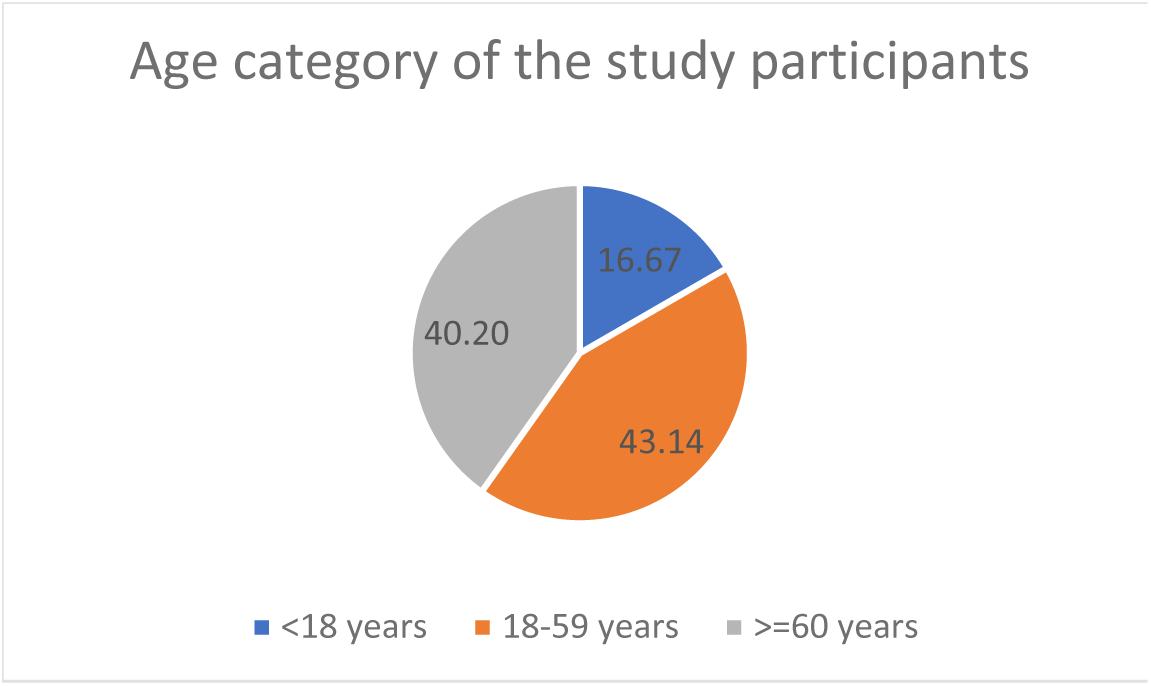
Age category of the study participants.

Among the study participants, 4.9% (n=5) had a history of prior documented infection with COVID-19 (Figure 2).

**Figure 2:**
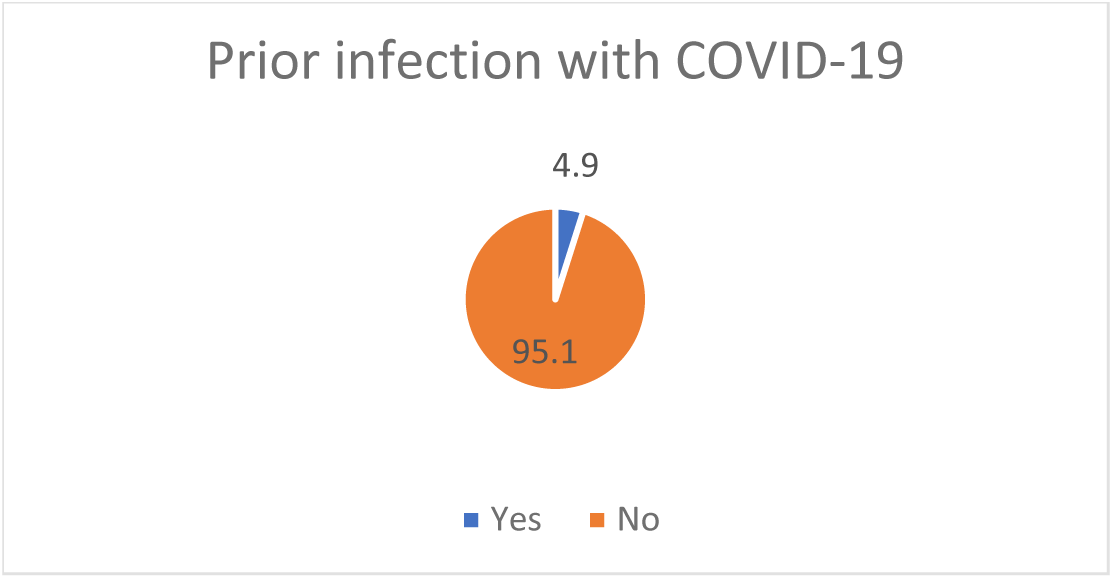
Proportion of participants with by prior infection with COVID-19.

The majority of the participants of the age group 18-59 years (56.82%) and those aged 60 years and greater (60.98%) were vaccinated. No participants below 18 years of age were vaccinated (Figure 3).

**Figure 3:**
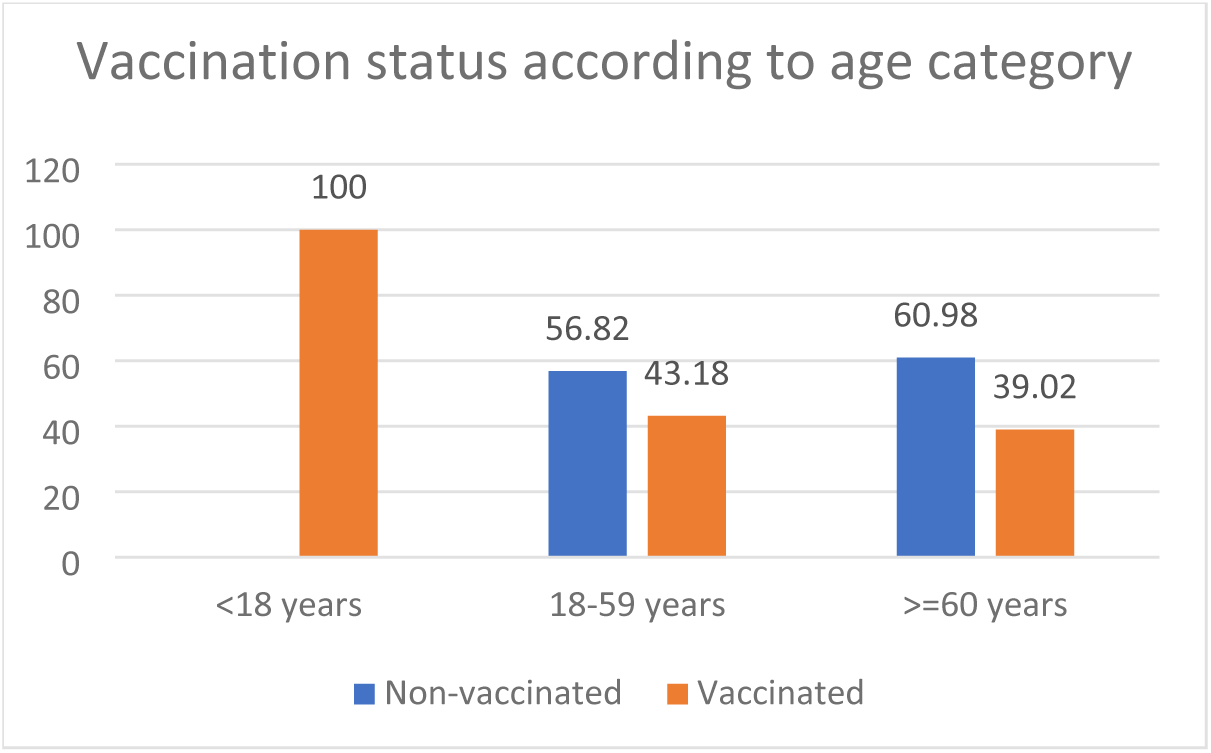
Vaccination status according to age category.

Among the study participants, 49% (n=50) had received at least one dose of a COVID-19 vaccine while 51% (n=52) had not. Among the vaccinated participants, the majority of them (46%) had received AstraZeneca (ChAdOx1-S [recombinant]) vaccine followed by Sinopharm COVID-19 vaccine (Verocell) (34%) and Janssen Ad26.COV2.S COVID-19 vaccine (20%) (Figure 4).

**Figure 4:**
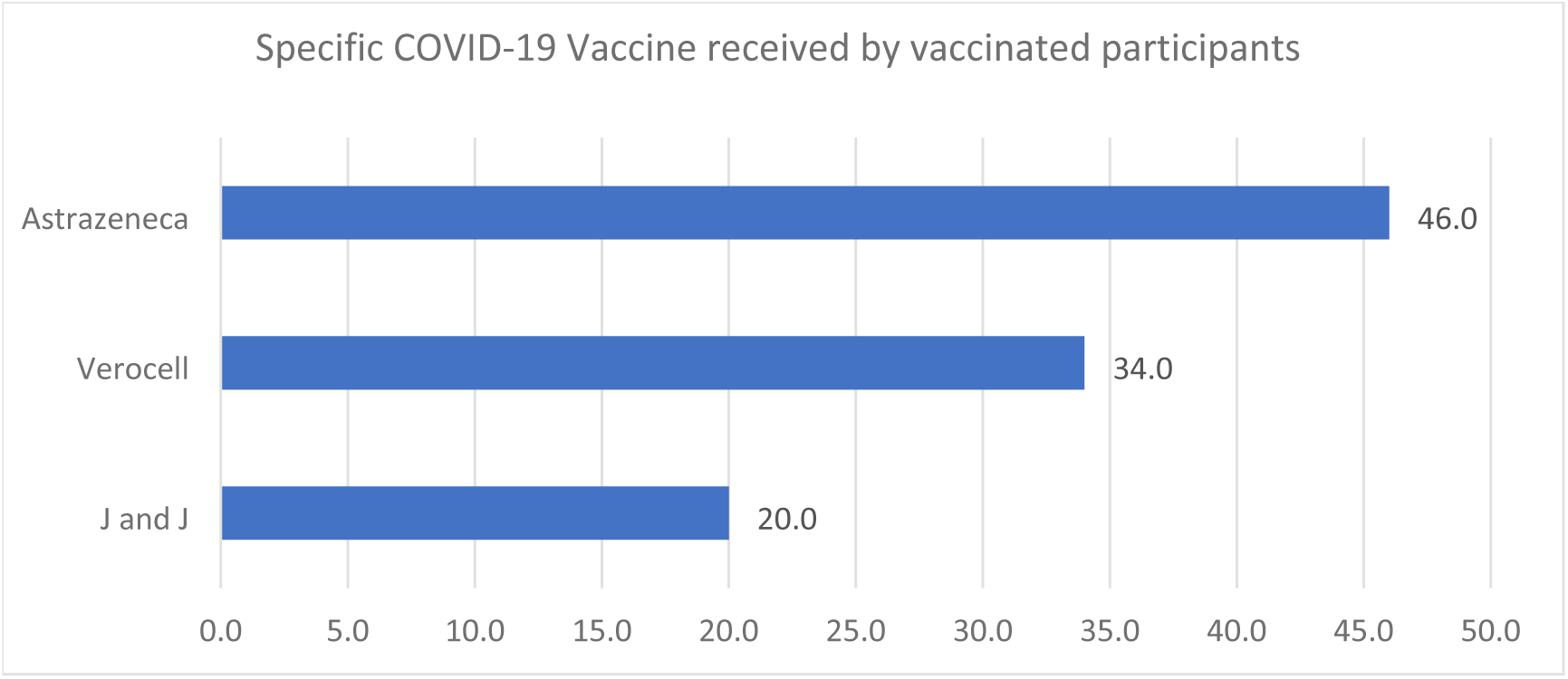
Specific type of COVID-19 vaccine received by the vaccinated participants.

Among the vaccinated participants, 50% (n=25) had received their last dose of vaccination within 3 months of admission and an equal percentage of participants had received their last dose greater than 3 months prior to admission (Figure 5).

**Figure 5:**
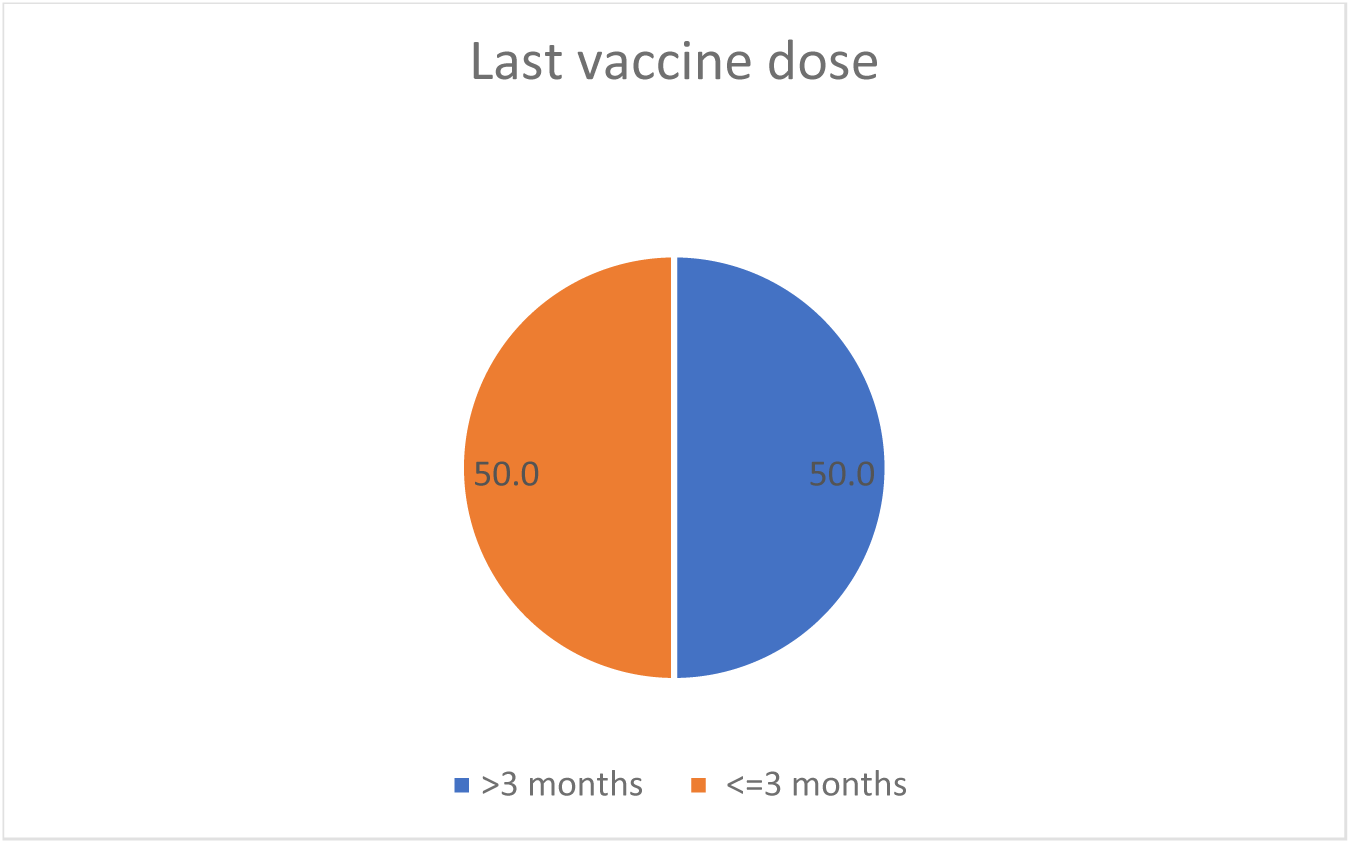
Proportion of the vaccinated participants by the timing of last dosing of the vaccine.

Among the vaccinated participants, 46% (n=23) had received only a single dose, while 54% (n=27) had received 2 doses of the vaccines (Figure 6). All of the participants who had received Janssen Ad26.COV2.S COVID-19 vaccine had received only one dose whereas 69.57% (n=16) of the participants receiving the AstraZeneca (ChAdOx1-S [recombinant]) vaccine (n=23) and 64.71% (n=11) of the participants receiving Sinopharm COVID-19 vaccine (Verocell) had received two doses of the vaccine (Figure 7).

**Figure 6:**
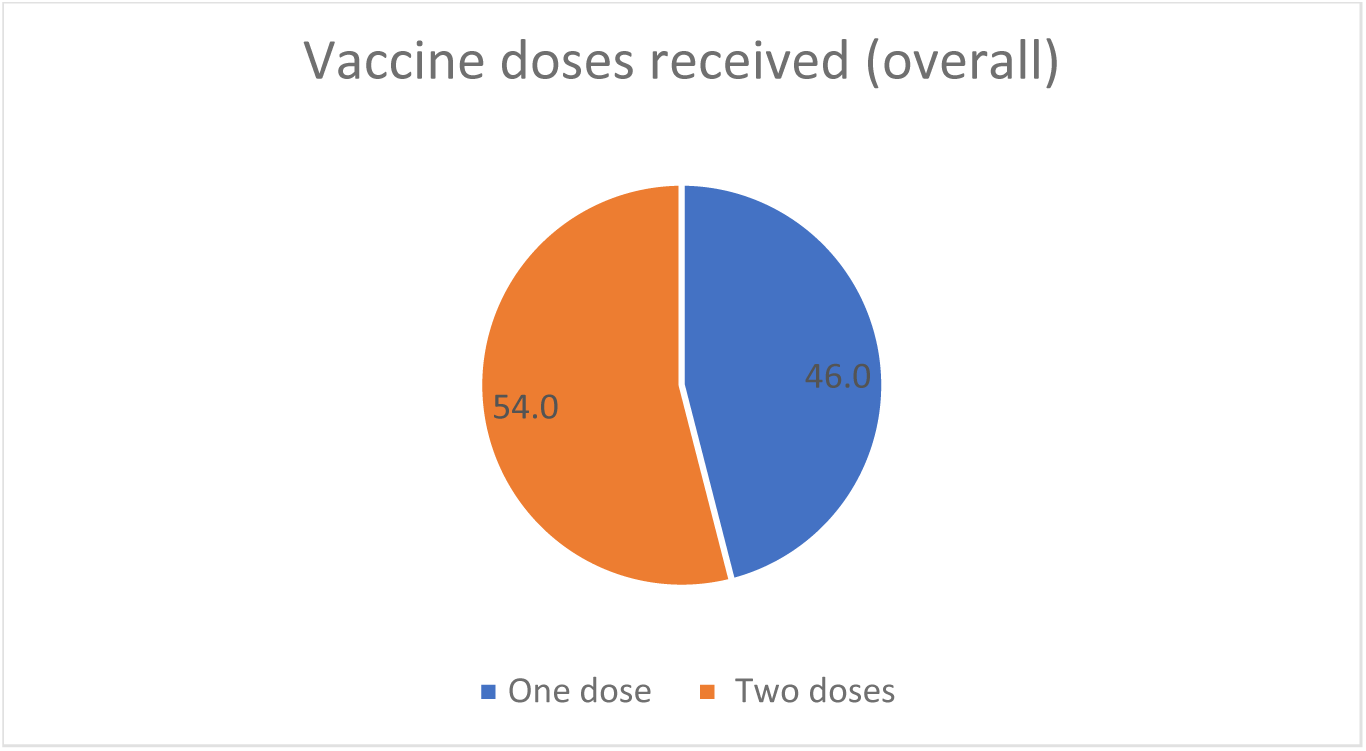
Proportion of study participants by the number of the doses of vaccine received (overall)

**Figure 7:**
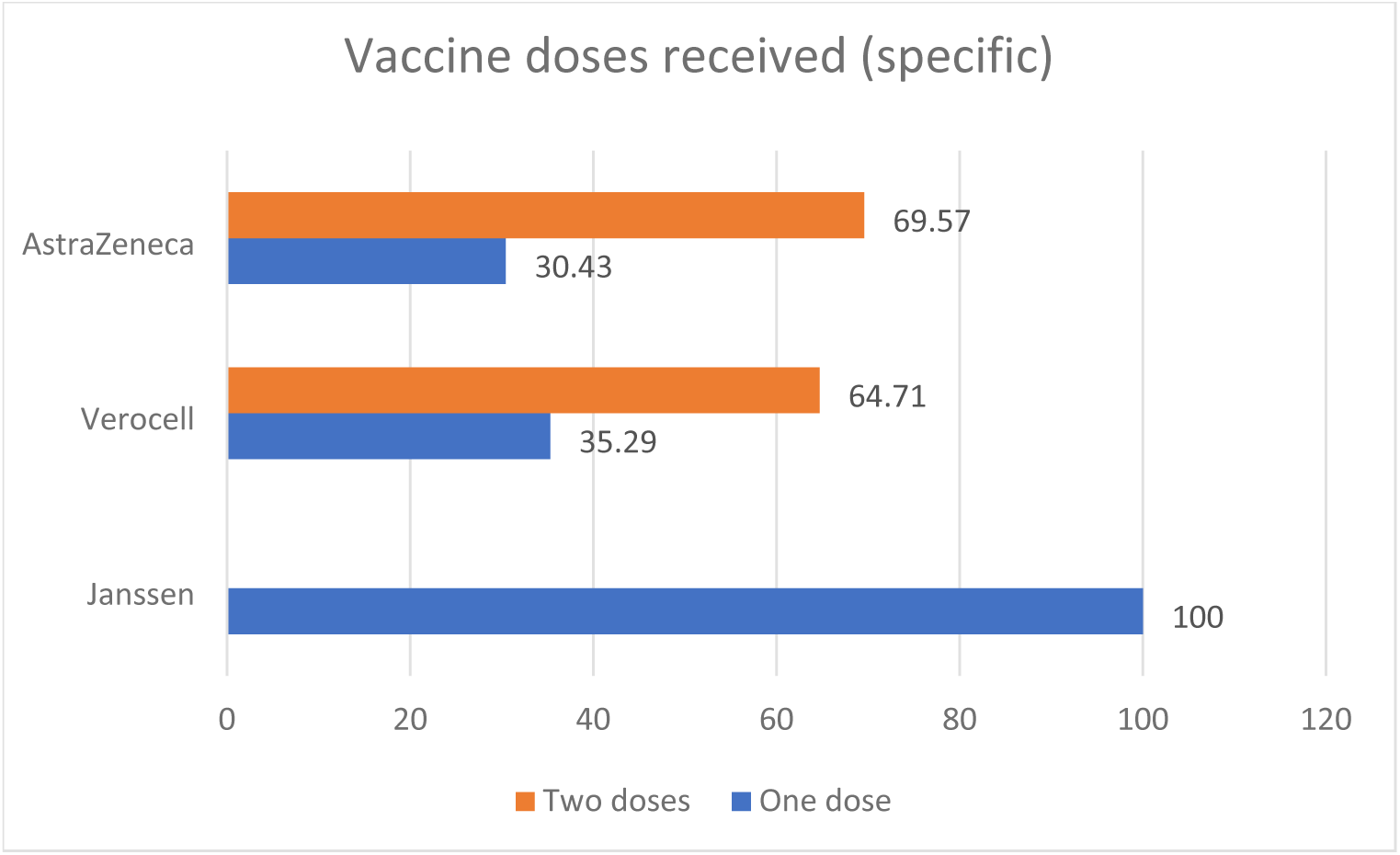
Proportion of study participants by the number of the doses of vaccine received (specific type of vaccine)

When the participants received two doses of the vaccine, the majority of them had not followed the dosing interval recommended by WHO. Only 11.76% (n=2) of participants receiving the Sinopharm COVID-19 vaccine (Verocell) and 17.39% (n=4) receiving the AstraZeneca (ChAdOx1-S [recombinant] vaccine) had adhered to the dosing interval recommended by the World Health Organization (WHO) (Table 1). Overall, only 22.22% of participants (n=6) had received the vaccines as per the WHO recommended interval, while 77.78% of participants (n=21) had not (Figure 8).

**Figure 8:**
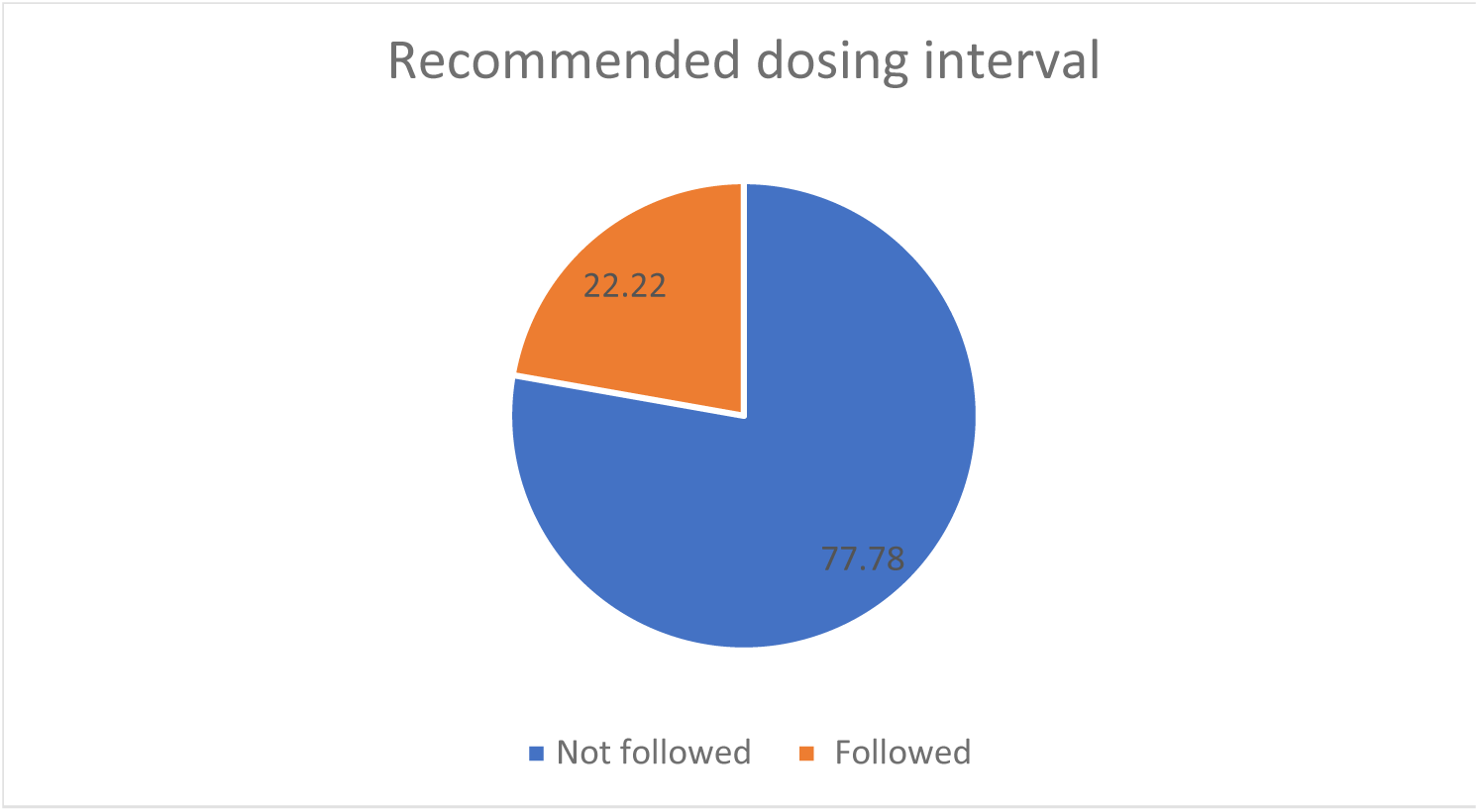
Proportion of study participants who received two vaccine doses by the adherence to recommended dosing interval.

**Table 1:**
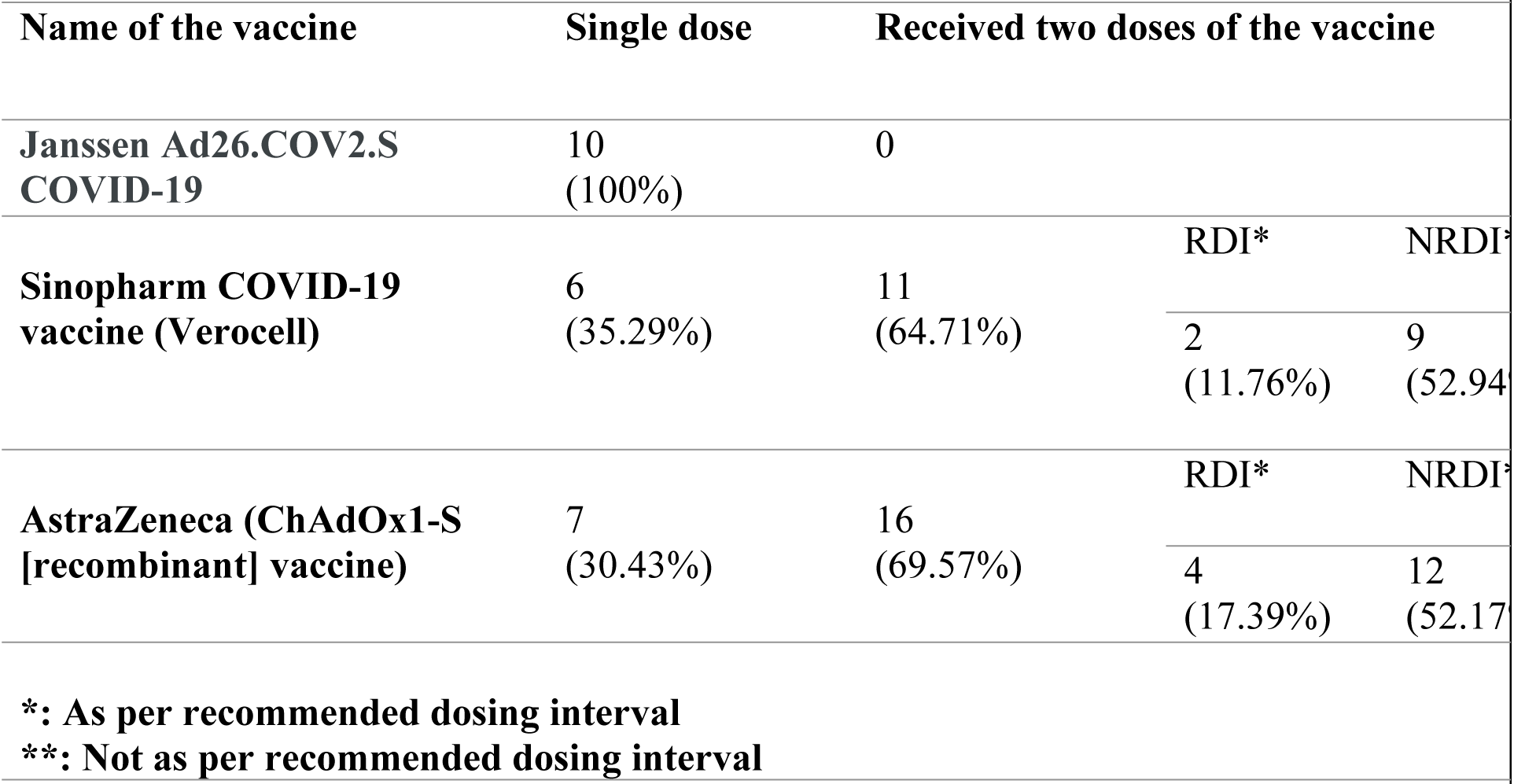
Patients who received one or two doses of the vaccines.

### Association Analysis

Vaccinated participants (n=50) had a significantly lower rate of severe disease compared to non-vaccinated participants (n=52) (20.00% vaccinated vs. 50.00% non-vaccinated, χ²=10.05, p=0.002) (Figure 9).

**Figure 9:**
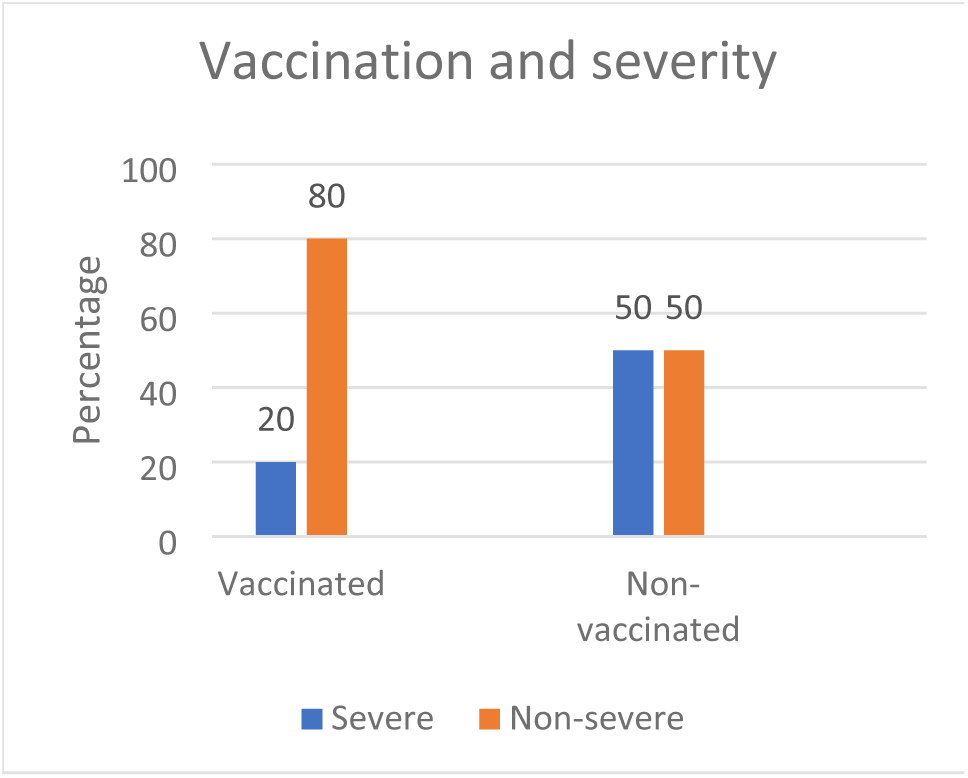
Vaccination status and severity of infection.

We examined the outcomes of participants in terms of recovery and mortality rates in our study. Among the vaccinated participants (n=50), 44 (88.00%) recovered, while 6 (12.00%) expired. Similarly, among the unvaccinated individuals (n=52), 42 (80.77%) recovered, and 10 (19.23%) expired (Table 2, Figure 10). No statistically significant difference was observed in recovery and mortality rates between the vaccinated and unvaccinated groups (χ²=1.008, p=0.315).

**Figure 10:**
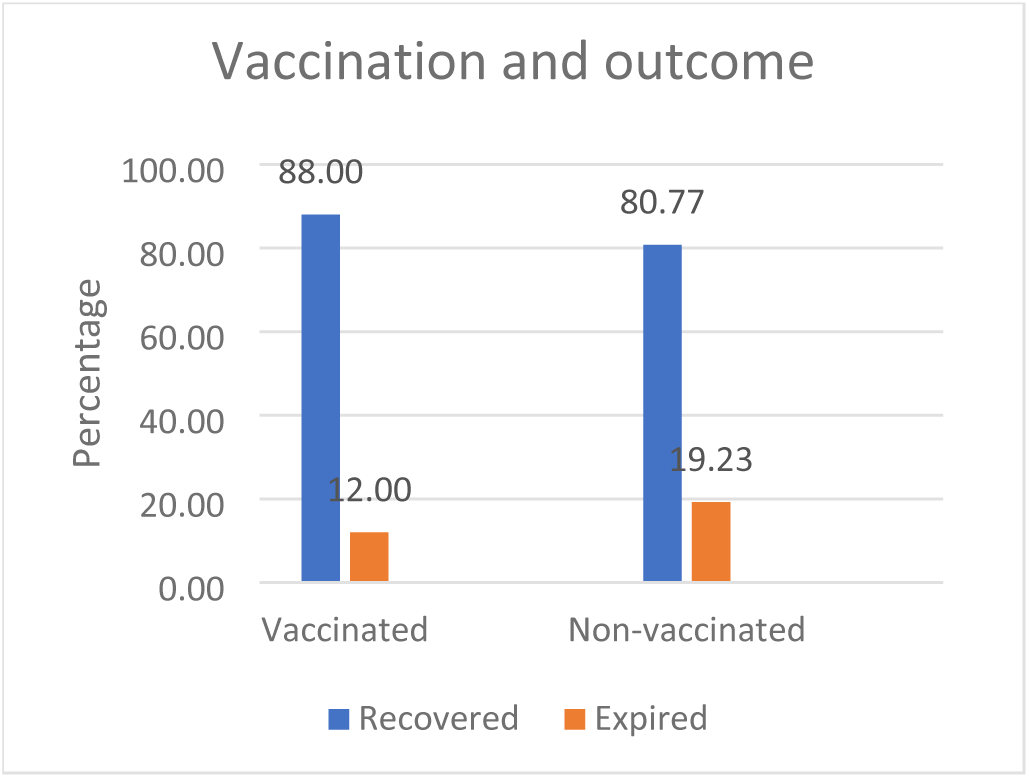
Vaccination status and outcome.

**Table 2:**
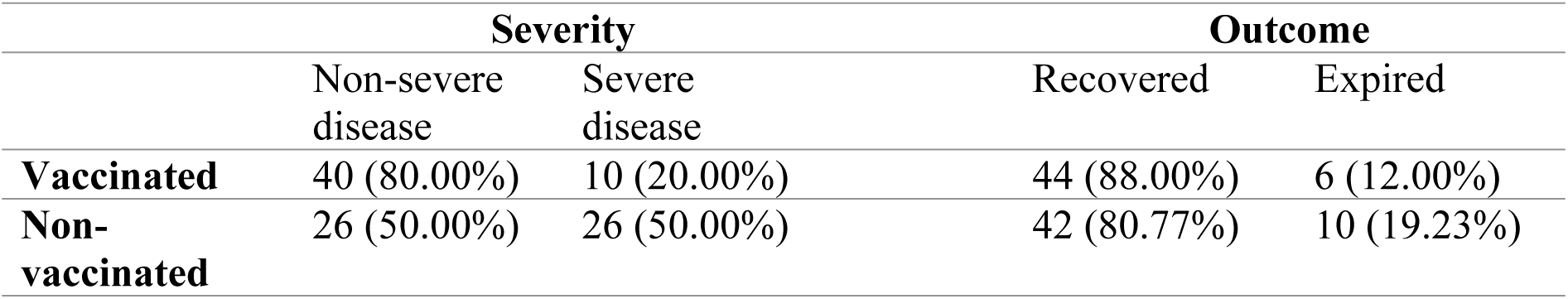
Outcomes among the vaccinated and non-vaccinated patients.

We analyzed the association between severity and vaccination status as well as between outcome and vaccination status among patients who received different types of vaccines (Table 3). There was no significant difference between severe and non-severe disease in the participants who received different types of vaccines (Fisher’s exact test, p = 0.296). Also, there was no significant association between different vaccine types and outcome (Fisher’s exact test, p = 0.740)

**Table 3:**
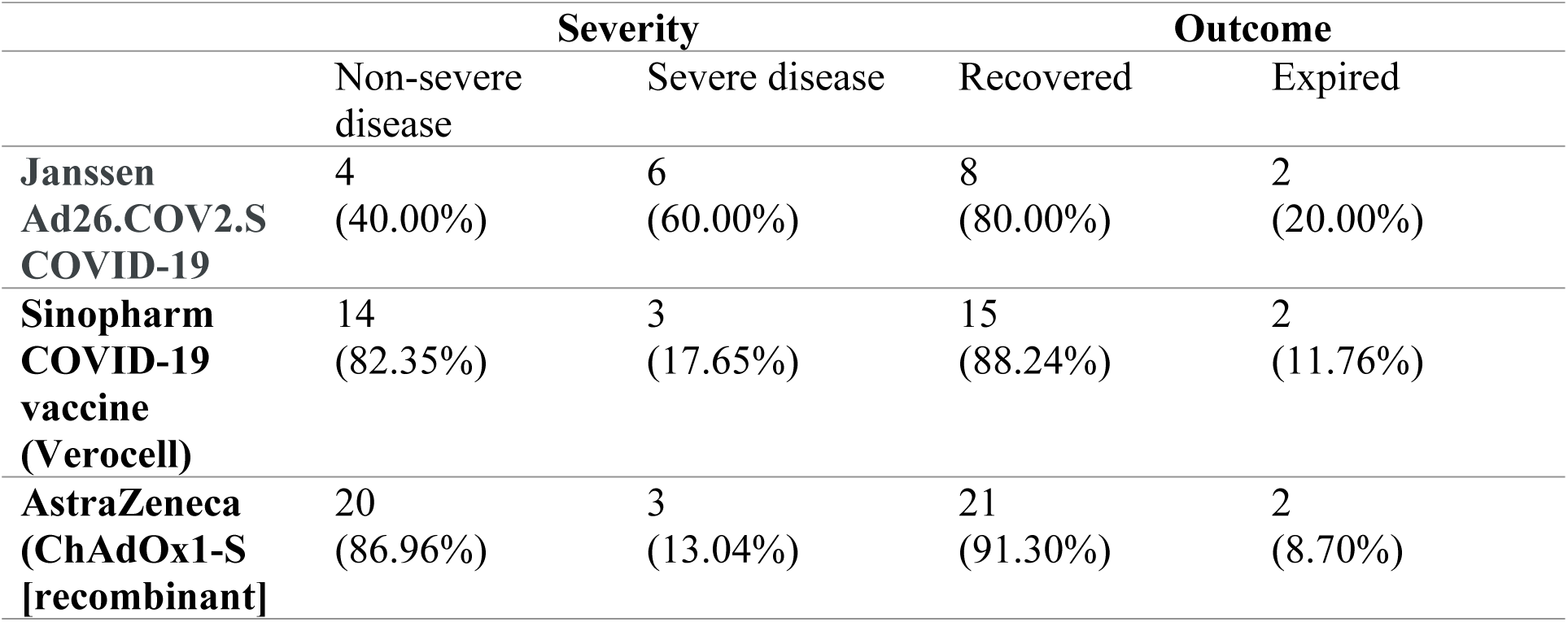
Association between different types of vaccines with severity and mortality of disease.

Among the vaccinated participants receiving two doses of the vaccine, we analyzed the association between adherence to recommended dosing intervals and severity of infection or mortality (Table 4). There were no statistically significant associations between participants who adhered to the recommended dosing interval and who did not in terms of severity (p = 0.56) or mortality (p = 1.00).

**Table 4:** Severity and outcome among the patients receiving two doses of vaccines who were and were not vaccinated with dosing intervals as per WHO recommendation.

|  | Severity |  | Outcome |  |
| --- | --- | --- | --- | --- |
|  | Non-severe disease | Severe disease | Recovered | Expired |
| **Recommended dosing interval** | 6<br>(100%) | 0 | 6<br>(100%) | 0 |
| **Not as per recommended dosing interval** | 16<br>(76.19%) | 5<br>(23.81%) | 18<br>(85.71%) | 3<br>(14.29%) |

Participants who received their last dose of vaccination within 3 months of admission (n=25) had a significantly lower rate of severe disease compared to those who received their last vaccine dose more than 3 months prior (8% vaccinated within 3 months vs. 24% vaccinated more than 3 months, χ²=4.500, p=0.034). Additionally, all participants who received their last vaccine dose within 3 months recovered, while 24% of those who received their last dose more than 3 months ago expired (Table 5). The association was found to be statistically significant (p=0.022, Fisher’s exact test).

**Table 5:** Severity and outcome among patients receiving last dose of their vaccine within 3 months and greater than 3 months.

| Last vaccination | Severity |  | Outcome |  |
| --- | --- | --- | --- | --- |
|  | Non-severe disease | Severe disease | Recovered | Expired |
| **<=3 months** | 23 (92.00%) | 2 (8.00%) | 25 (100.00%) | 0 |
| **>3 months** | 17 (68.00%) | 8 (32.00%) | 19 (76.00%) | 6 (24.00%) |

We stratified the severity and outcome in vaccinated and non-vaccinated participants in terms of whether or not they had any comorbidities (Table 6). The major co-morbidities noted in our study participants were hypertension, diabetes mellitus, and chronic obstructive pulmonary disease.

**Table 6:** Severity of infection and outcome among vaccinated and non-vaccinated patients.

|  |  | Severity |  | Outcome |  |
| --- | --- | --- | --- | --- | --- |
|  |  | Non-severe disease | Severe disease | Recovered | Expired |
| **Comorbidities absent** | Vaccinated | 14 (87.50%) | 2 (12.50%) | 15 (73.75%) | 1 (6.25%) |
|  | Non-vaccinated | 25 (52.08%) | 23 (47.92%) | 41 (85.42%) | 7 (14.58%) |
| **Comorbidities present** | Vaccinated | 26 (76.47%) | 8 (23.53%) | 29 (85.29%) | 5 (14.71%) |
|  | Non-vaccinated | 1 (25.00%) | 3 (75.00%) | 1 (25.00%) | 3 (75.00%) |

Vaccinated participants with or without comorbidities had significantly lower rates of severe disease compared to non-vaccinated participants (Fisher’s exact test, p = 0.065 for participants without comorbidities; χ²=6.323, p = 0.017 for participants with comorbidities).

Vaccinated participants with comorbidities had significantly lower mortality rate (14.71%) compared to non-vaccinated participants (75.00%), indicating an approximately 5-fold higher mortality rate among the non-vaccinated group (Fisher’s exact test, p=0.024).

With comorbidities absent, we found no statistical significance through Fisher’s exact test (p=0.667) between the mortality rates of vaccinated and unvaccinated groups.

Also, we stratified the severity of infections and outcomes according to age factor (Table 7). Statistical analysis showed no significant difference between severity and vaccination status among participants aged 60 years and above (Fisher’s exact test, p = 0.287). But there was a significant association between severity and vaccination status among the participants of the age group 18-59 years (χ²=8.481, p = 0.004). There was no significant difference in outcome and vaccination status among the participants of the age group 18-59 years (Fisher’s exact test, p = 0.210) and age group >=60 years (Fisher’s exact test, p = 1.00)

**Table 7:**
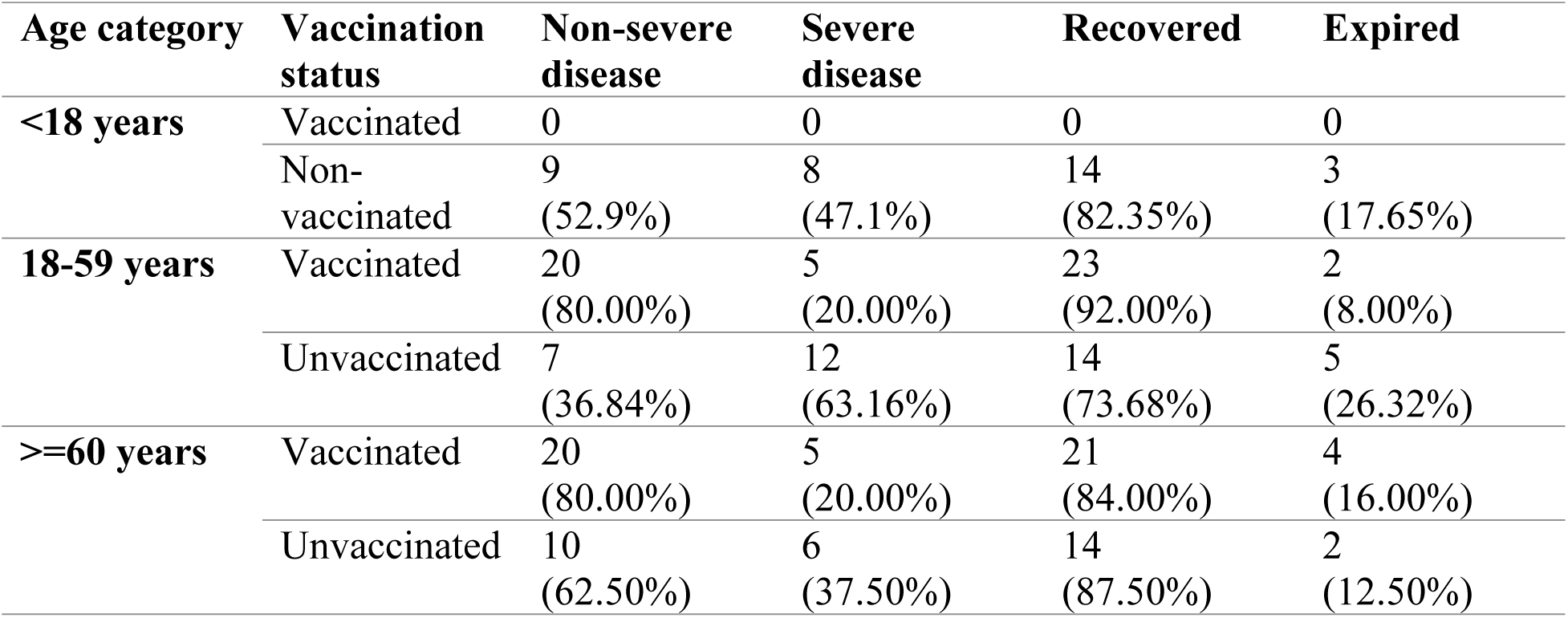
Severity and outcome of COVID-19 infections among vaccinated and non-vaccinated groups according to different age category.

| Age category | Vaccination status | Non-severe disease | Severe disease | Recovered | Expired |
| --- | --- | --- | --- | --- | --- |
| <18 years | Vaccinated | 0 | 0 | 0 | 0 |
|  | Non-vaccinated | 9<br>(52.9%) | 8<br>(47.1%) | 14<br>(82.35%) | 3<br>(17.65%) |
| 18-59 years | Vaccinated | 20<br>(80.00%) | 5<br>(20.00%) | 23<br>(92.00%) | 2<br>(8.00%) |
|  | Unvaccinated | 7<br>(36.84%) | 12<br>(63.16%) | 14<br>(73.68%) | 5<br>(26.32%) |
| >=60 years | Vaccinated | 20<br>(80.00%) | 5<br>(20.00%) | 21<br>(84.00%) | 4<br>(16.00%) |
|  | Unvaccinated | 10<br>(62.50%) | 6<br>(37.50%) | 14<br>(87.50%) | 2<br>(12.50%) |

We analyzed the association between prior infection and severity as well as outcome of current infection (Table 8). Fisher’s exact test failed to reveal a significant association between prior infection and current infection severity (p = 0.158) or outcome (p = 1.0).

**Table 8:** Prior infection and severity or outcome of current infection.

|  | Severity |  | Outcome |  |
| --- | --- | --- | --- | --- |
|  | Non-Severe disease | Severe disease | Recovered | Expired |
| **Prior infection with COVID-19** | 5<br>(100%) | 0 | 5<br>(100%) | 0 |
| **No Prior infection with COVID-19** | 61<br>(62.89%) | 36<br>(37.11%) | 81<br>(83.51%) | 16<br>(16.49%) |

## Discussion

The purpose of this study was to look into the relationship between vaccination status, re-infection, and clinical outcomes among patients admitted to the COVID High Dependency Unit (HDU). We looked into things like the type of COVID-19 vaccine used, the number of vaccine doses given, adherence to recommended dosing intervals, time since vaccination, and the effect of vaccination on severity and outcome. Furthermore, we investigated the impact of comorbidities and prior infection on disease severity and outcome. We believe that this study will contribute to the understanding of COVID-19 vaccine effectiveness by guiding clinical decision-making and public health interventions. We compare our findings to the existing literature, identify limitations, and suggest future research directions.

The gender distribution among participants in our study was relatively balanced, with 52% male and 48% female. This gender distribution reflects our study population’s inclusion of both males and females, allowing for a more representative analysis. In terms of vaccination status, we found that 49% of participants had received at least one dose of COVID-19 vaccine, while 51% had not received any. This distribution sheds light on vaccination coverage in the study population, indicating that a sizable proportion of people had not yet received any COVID-19 vaccine at that time. This finding may have implications for how the results of the study on the relationship between vaccination status and the severity or outcome of the current infection are interpreted.

Our finding showed that vaccination with the COVID-19 vaccine is associated with a significantly lower risk of severe disease is consistent with the studies in the past which investigated the association between vaccination and disease severity.^13–15^ However, unlike these studies, our study failed to demonstrate a significant relationship between vaccination status and the outcome (recovery or death).

One study by Bernal et al. showed that one dose of either BNT162b2 or ChAdOx1-S offered protection against severe disease for > 6 weeks. Our study finding is in line with this finding as it showed that the vaccinated patients who were infected with COVID-19 within 3 months of vaccination had a significantly lower risk of severe disease and mortality.^16^

Our study had some limitations. First, a relatively small sample size in our study may limit the generalizability of the findings to broader populations. Second, the study design was cross-sectional, which limits our ability to establish causal relationships. Longitudinal studies or randomized controlled trials would provide stronger evidence of the impact of vaccination and comorbidities on disease outcomes. Additionally, there may have been confounding factors that were not accounted for in our analysis, such as socioeconomic status or access to healthcare. These factors could potentially influence both vaccination status and disease outcomes.

Lastly, our study focused on a specific population or setting, which may limit the generalizability of the findings to other regions or healthcare settings. Variations in vaccination rates, healthcare infrastructure, and population characteristics could affect the observed associations.

Despite these limitations, our study contributes valuable insights into the association between vaccination status, comorbidities, and disease outcomes in COVID-19 patients. Further research with larger sample sizes, more rigorous study designs, and diverse populations is warranted to build upon our findings and provide a more comprehensive understanding of these relationships.

## Conclusion

Our findings suggest that COVID-19 vaccination is associated with a reduced risk of severe disease among individuals with or without comorbidities and decreased risk of mortality among those with comorbidities. However, larger studies are needed to validate these findings and further explore the impact of vaccination on disease outcomes. These findings support the ongoing efforts to promote COVID-19 vaccination as a crucial public health intervention.

## ACKNOWLEDGEMENT

We thank all the patients participating in this study.

## ETHICAL APPROVAL

The study protocol (Code no. 48/2022 P) was approved by Nepal Health Research Council.

## CONSENT FOR PUBLICATION

Written informed consent was obtained from the patients for publication of this original article. A copy of the written consent is available for review by the Editor-in-Chief of this journal on request.

## AUTHORS CONTRIBUTION

All the authors contributed equally to writing and preparing the manuscript. The final version of the article is approved by all authors.

## RESEARCH REGISTRATION

No new surgical techniques or new equipment/technology was used.

## PROVENANCE AND PEER REVIEW

Not commissioned, externally peer reviewed.

## DATA AVAILABILITY STATEMENT

Datasets generated during and/or analyzed during the current study are available upon reasonable request.

